# Mapping Long COVID: Spatial and Social Inequities Across the United States

**DOI:** 10.1101/2025.08.21.25334183

**Authors:** Zhetao Chen, Bingnan Li, Yewen Chen, Jialing Liu, Fangzhi Luo, Kehinde Olawale Ogunyemi, Yang Ge, Yuan Ke, Yang Yang, Xianyan Chen, Ye Shen The National COVID Cohort Collaborative

## Abstract

**Background:** Long COVID affects a substantial portion of the U.S. population, yet its spatiotemporal distribution remains poorly characterized. The emergence of the Omicron variant and persistent sociodemographic disparities may contribute to regional variation in long COVID risk. Understanding the patterns of long COVID is essential to implementing targeted and equitable public health interventions.

**Methods:** This retrospective study utilized data from the National COVID Cohort Collaborative (N3C), covering 5,652,474 COVID-19 cases and 41,694 long COVID cases across 1,063 U.S. counties from 2021 to 2024. Temporal patterns of long COVID were analyzed before and after the Omicron variant’s emergence, and spatial patterns were assessed using Moran’s I and Getis statistics. Bayesian spatial random effect models were employed to evaluate the associations between long COVID incidence and sociodemographic factors such as economic vulnerability, healthcare access, and mobility.

**Findings:** Quarterly long COVID incidence ranged from 0.015% to 14.29%. Before the emergence of the Omicron variant, incidence was 204 cases per 10,000 COVID-19 cases, compared with 248 cases per 10,000 COVID-19 cases after Omicron emergence (p < 0.001). After Omicron’s emergence, 48.8% [328 of 673] of counties showed significant spatial correlation (p < 0.05), up from 43.5% [293 of 673] prior. High-risk areas became more concentrated in inland regions, while low-risk areas clustered along the East Coast. Long COVID incidence was significantly associated with economic vulnerability, limited healthcare access, and mobility constraints, with these sociodemographic disparities consistently driving its spatial disparities over time.

**Interpretation:** These findings underscore the need to address spatial and social inequities in long COVID risk. Targeted public health interventions, particularly in economically and geographically vulnerable regions, are essential to ensure equitable access to diagnosis, care, and resource allocation.

**Funding:** Y. Shen received partial support from NIH grants/contracts R35GM146612, R01AI170116, and 75N93019C00052.

## 1. Introduction

Long COVID, or post-acute sequelae of SARS-CoV-2 infection (PASC), has become a major public health concern due to its long-term impacts^1,2^. In 2022, approximately 6.9% of adults reported having ever experienced long COVID, and 3.4% were currently experiencing it, illustrating the substantial prevalence and persistence of this condition^3^. These long-term symptoms can involve multiple physiological systems, such as respiratory, neurological, cardiovascular, and gastrointestinal, leading to fatigue, cognitive impairment, shortness of breath, and other chronic issues^4^. Although many epidemiological studies have explored individual-level risk factors for long COVID-19^5,6^, our understanding of its spatial and temporal incidence dynamics remains limited.

The distribution of long COVID exhibits complex spatial and temporal disparities. The spread of acute COVID-19 was highly uneven across regions^7^, and emerging evidence suggests that long COVID prevalence also varies geographically^8,9^. According to a June 2022 survey conducted by the Centers for Disease Control and Prevention (CDC), the prevalence of current long COVID symptoms among the United States (U.S.) adults varied substantially across states, ranging from 4.5% in Hawaii to 12.7% in Kentucky^10^. Additionally, temporal shifts have also been observed, particularly before and after the emergence of the Omicron variant^11^. Beyond these patterns, socioeconomic and demographic factors can play a crucial role in accounting for long COVID risk^10,12^. Prior studies have shown that prevalence differs across age groups and racial/ethnic populations^13^. However, previous studies have rarely examined the spatiotemporal patterns of long COVID incidence and how these factors dynamically influence spatial disparities in long COVID incidence.

In this study, we analyzed county-level long COVID data from the National COVID Cohort Collaborative (N3C) and sociodemographic data from the Social Vulnerability Index (SVI), covering 1,063 U.S. counties over the period 2021–2024. Our analysis advances prior work by providing a detailed spatiotemporal assessment and evaluating the influence of community-level social factors on incidence patterns. These findings can help guide targeted public health strategies and inform equitable healthcare resource allocation.

## 2. Methods

### 2.1 Study population and data collection

We constructed two datasets from the N3C database (version 185), one for acute COVID-19 and one for long COVID, based on daily EHR data from July 1, 2020 to March 31, 2024 (**Figure** S1 of the Supplementary Material). The data included individual-level records of confirmed cases, ZIP codes of site locations, and associated demographics, contributed by 62 sites across U.S. counties. Due to the sparse spatial distribution of sites, this study focuses on county-level analysis rather than site-level analysis to improve the robustness of the results. By summary, 1,063 of 3,144 counties with at least one reported long COVID case were included in the incidence analysis. Moreover, to enable valid pre-post comparisons around the Omicron-dominant period, we further restricted the related comparison analysis to 673 counties with available long COVID data both before and after this period.

To understand factors potentially influencing long COVID incidence, we incorporated socioeconomic and demographic variables from SVI, developed by the Centers for Disease Control and Prevention and the Agency for Toxic Substances and Disease Registry^14^. We used two versions of the SVI data to align with our pre- and post-Omicron analyses: the 2022 SVI, based on the 5-year (2018–2022) American Community Survey (ACS), was used for the post-Omicron period, and the 2020 SVI, based on the 5-year (2016–2020) ACS, was used for the pre-Omicron period. This approach ensures that the socioeconomic context used in each analysis reflects the corresponding time frame as accurately as possible, which is particularly important given potential shifts in community characteristics over the course of the pandemic. SVI was designed to identify communities that may need support before, during, or after public health emergencies. In this study, nine SVI-related variables, along with nine demographic and geographic factors, were analyzed. In particular, these SVI variables reflected socioeconomic status, household characteristics, racial and ethnic minority status, housing type, and access to transportation^15^.

### 2.2 Study outcomes

The incidence of long COVID was used as the outcome of interest. To assess evolution of long COVID incidences over time, we employed dynamic incidence risk calculation method (**Figures** S2-S3 of the Supplementary Material). By comparing long COVID EHR cases on specific time periods across various spatial regions with the corresponding at-risk acute COVID-19 populations, we derived two outcomes for the long COVID incidence. One was the incidence before and after the Omicron-dominant period (i.e., January 2022), which was set based on the standard provided by the WHO^16^. The other outcome was the quarterly incidence computed using three-month intervals based on natural calendar months given emerging evidence on the seasonal variability of COVID-19^17^.

### 2.3 Statistical analysis

To assess spatial local patterns in long COVID incidence across U.S. counties, we employed both local Moran’s I and Getis statistics^18,19^ (**Figure** S4 of the Supplementary Material). Local Moran’s I quantifies the degree of similarity or dissimilarity between a county and its neighbors, with positive values indicating similarity and negative values indicating dissimilarity. In contrast, Getis identifies statistically significant spatial clusters of high (hot spots) or low (cold spots) incidence.

To accurately estimate potential relationships between variables and long COVID incidence, simple stepwise regression methods were used to identify a subset of variables that best explained the variation in long COVID incidence. Given the right-skewed distribution of long COVID incidence, we applied a logarithmic transformation to stabilize variance and improve model performance (**Figure** S8 of the Supplementary Material). Based on the selected variables, spatial random effects models were then employed to account for spatial heterogeneity. The model can be written as:

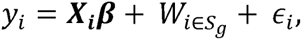

where *y_i_* is the incidence of the *i*-th county, ***X*** is the covariables selected via stepwise regression models. To avoid confounding introduced by spatially correlated random effects, we modeled the zero-mean Gaussian spatial random effect 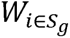 as subregion-level independent errors, with variances 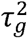 varying across subregions for *g* = 1, …,5(**Figure** S5 of the Supplementary Material). The term *ε_i_* represents zero-mean Gaussian errors with the variance of *τ*^2^. To avoid potential multicollinearity among covariables, under a Bayesian setting, we used a ridge-like prior for the coefficients ***β*** to stabilize estimates, specified as zero-mean Gaussian with a small variance of 0.1. All analyses were conducted using R statistical software (version 4.4.0)^20^.

## 3. Results

### 3.1 Overall description

From 2020 to 2024, a total of 5,652,474 COVID-19 cases were recorded in the N3C database, among which 41,694 long COVID cases were identified based on the dynamic incidence risk described in **Figure** S2, resulting in an overall incidence risk of 0.74% across the study period. The quarterly incidence of long COVID ranged from 0.015% to 14.29%, with 66.0% of counties recording incidence rates in the interval [0.015%, 1.50%] (**Figure** 1a). The average incidence across counties peaked in the fourth quarter of 2021 (October-December) at 2.07%, followed by another high in the first quarter of 2022 (January-March) at 1.91%, before declining thereafter (**Figure** 1b). These two peaks coincided with the period when the Omicron variant rapidly replaced Delta as the dominant strain in the U.S. during 2022 Q1. A Wilcoxon signed-rank test (p < 0.0001) indicated a statistically significant difference in long COVID incidence before and after the Omicron-dominant period, suggesting a shift in the trend associated with the emergence of Omicron.

**Figure 1.**
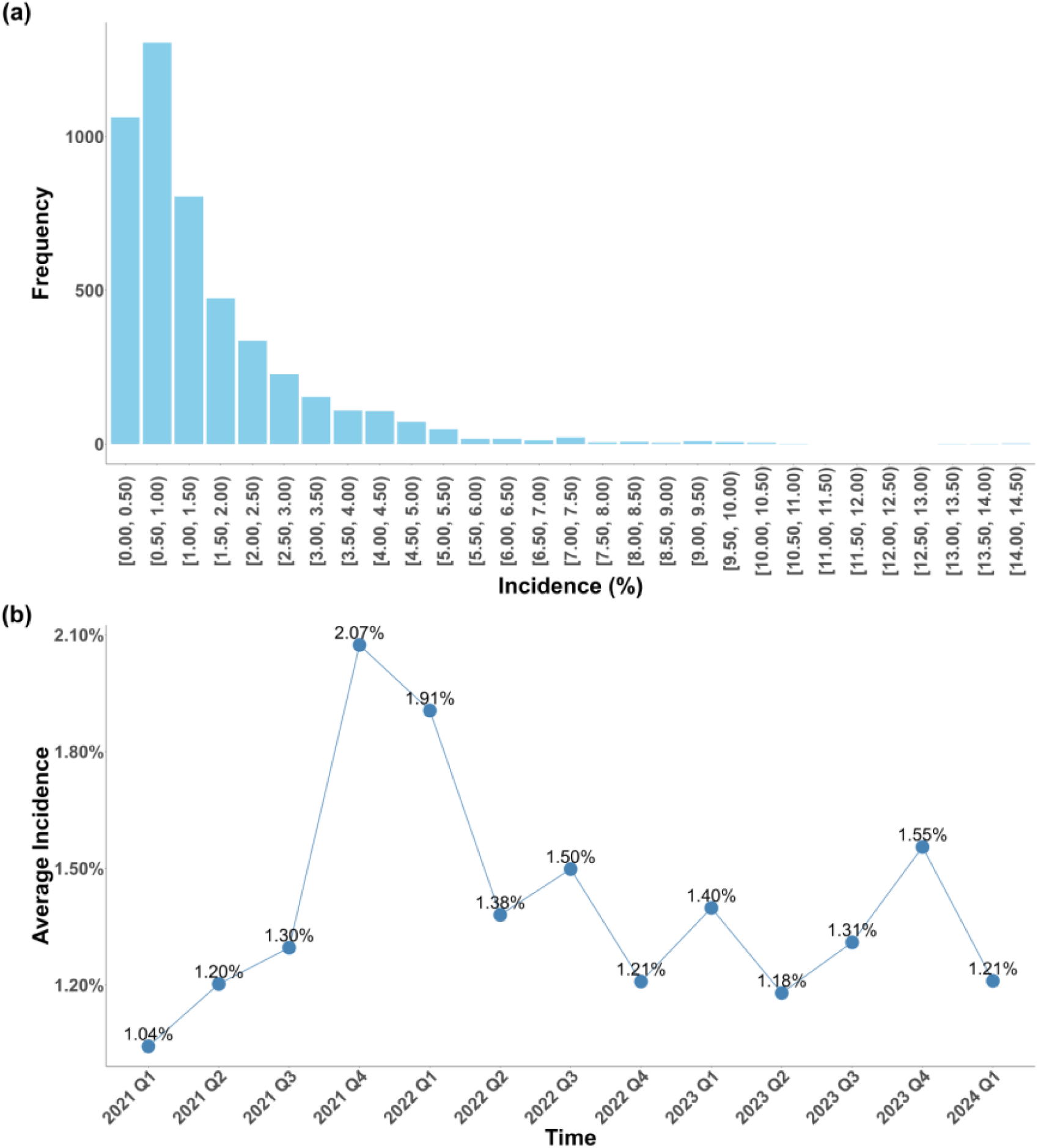
Long COVID incidences. (a) Its distribution in each interval; and (b) Time-trend of average incidence across counties from January 2021 to March 2024.

To facilitate geographic comparisons, we adopted a commonly used five-region classification of the U.S.^21^(**Figure** S5). All five U.S. regions exhibited a substantial increase in long COVID incidence following the Omicron surge (**Table** S1 of the Supplementary Material). Specifically, more than two-thirds of counties in each region experienced an increase with the Northeast showing the highest proportion (75%) and the South showing the lowest (66.67%). Despite regional variation, the overall proportion of counties with increased incidence was moderately high at 68.95%, highlighting the significant epidemiologic shift associated with the emergence of Omicron. In detail, the patterns of increasing or decreasing county-level incidence before and after the Omicron-dominant period exhibited local spatial similarity (**Figure** S6 of the Supplementary Material). Moreover, the incidence varied notably across regions (**Table** S2 of the Supplementary Material). The West consistently exhibited the highest average incidence, increasing from 2.42% before Omicron to 2.86% after its emergence. The Midwest and Northeast also experienced considerable increases, with post-Omicron incidences reaching 2.79% and 2.45%, respectively. Interestingly, while the South maintained a moderate level of incidence, increasing from 2.21% to 2.53%, the Southwest was the only region where the average incidence slightly declined after the Omicron surge, from 1.85% to 1.78%.

### 3.2 Spatial correlation analysis

Local Moran’s I analysis revealed that the total proportion of counties with significant correlations increased from 43.5% before the emergence of Omicron to 48.8% afterward (**Table** S3 of the Supplementary Material). Positive spatial autocorrelation was more prevalent than negative in both periods. Specifically, the proportion of counties showing positive correlations rose from 32.7% to 35.4%, while the proportion with negative correlations increased from 10.8% to 13.4%. Regionally, the proportion of counties with positive correlations decreased in the Northeast (from 43.3% to 21.7%) and South (from 51.6% to 50.8%), remained relatively stable in the Midwest (from 22.8% to 23.2%), and increased notably in the West (from 11.1% to 32.2%) and Southwest (from 0.0% to 38.2%). For negative correlations, the proportion declined slightly in the South (from 14.7% to 14.3%), Northeast (from 8.3% to 5.0%), and Southwest (from 11.8% to 8.8%), but increased in the Midwest (from 7.2% to 12.7%) and West (from 11.1% to 20.0%). Overall, counties with significant spatial correlations were primarily located in the Middle and eastern regions (**Figure** S7 of the Supplementary Material).

### 3.3 High- and low-risk areas

Getis statistics identified notable shifts in the spatial distribution of both high-risk and low-risk areas for long COVID incidence before and after the dominance of the Omicron variant (**Table** 1). The total proportion of counties classified as high-risk increased markedly from 14.9% to 27.2%. Regionally, the Midwest experienced the largest increase in high-risk counties, rising from 16.9% to 35.9%, followed by the West (22.2% to 40.0%) and the South (14.7% to 23.8%). In contrast, the Northeast had no high-risk counties identified in either period, and the Southwest showed a slight decrease from 8.8% to 5.9%. Conversely, the total proportion of counties identified as low-risk declined from 28.7% to 21.5%. The South and Northeast showed decreases in low-risk counties – from 51.6% to 41.3% and 51.7% to 26.7%, respectively. The Midwest saw a complete drop from 13.1% to 0.0%, while the West increased slightly from 0.0% to 12.2%. Notably, the Southwest showed a substantial rise in low-risk counties, from 2.9% to 41.2%. Overall, the results indicate an expansion of high-risk clusters – especially across the Midwest and West – after the emergence of the Omicron variant, whereas low-risk areas became more concentrated in the Southwest, as visualized in **Figure** 2.

**Table 1.** Changes in the number and proportion of counties with high and low long COVID incidence risk before and after Omicron dominance across different subregions of the United States (673 counties).

| Region | The number (proportion<sup>a</sup>) of |  | The number (proportion) of |  |
| --- | --- | --- | --- | --- |
|  | counties in high risk |  | counties in low risk |  |
|  | Before | After | Before | After |
| South | 37 (14.7%) | 60 (23.8%) | 130 (51.6%) | 104 (41.3%) |
| Northeast | 0 (0.0%) | 0 (0.0%) | 31 (51.7%) | 16 (26.7%) |
| Midwest | 40 (16.9%) | 85 (35.9%) | 31 (13.1%) | 0 (0.0%) |
| West | 20 (22.2%) | 36 (40.0%) | 0 (0.0%) | 11 (12.2%) |
| Southwest | 3 (8.8%) | 2 (5.9%) | 1 (2.9%) | 14 (41.2%) |
| Total | 100 (14.9%) | 183 (27.2%) | 193 (28.7%) | 145 (21.5%) |

<sup>a</sup> Proportion = (Number of high- or low-risk counties) / (Total number of counties in the region)

**Figure 2.**
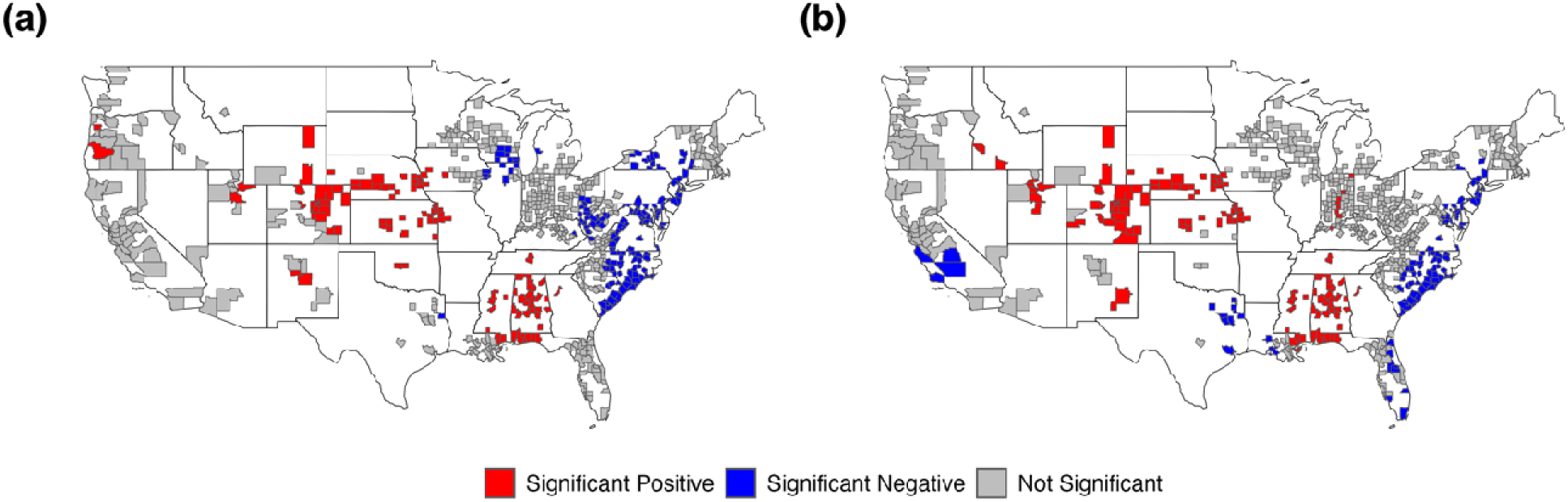
Spatial clustering of long COVID incidence across U.S. counties based on Getis statistic before (a) and after (b) Omicron dominance.

### 3.4 Associations between social vulnerability factors and long COVID incidence

Stepwise regression identified different sets of variables for the periods before and after Omicron dominance (**Table** S4 and S5 of the Supplementary Material). The spatial random effect models were then developed using the selected variables. **Figure** 3 presents the results for variables that were selected in both periods, while variables that differed between the two periods are shown in **Figure** S10 of the Supplementary Material. Urban counties consistently exhibited lower long COVID incidence than rural counties in both the pre- and post-Omicron periods (–0.181, 95% CI: –0.292 to –0.070 before; –0.123, –0.230 to –0.016 after). Counties with a higher proportion of adults without a high school diploma also showed significant negative associations across both periods (–0.018, –0.033 to –0.003 before; –0.038, –0.055 to –0.021 after), as did those with a higher proportion of racial and ethnic minority populations (–0.005, –0.009 to –0.001 before; – 0.010, –0.014 to –0.007 after). In contrast, a higher proportion of disabled residents was significantly positively associated with long COVID incidence only before Omicron (0.047, 0.030 to 0.063), with a weaker but still significant association observed afterward (0.027, 0.007 to 0.046).

**Figure 3.**
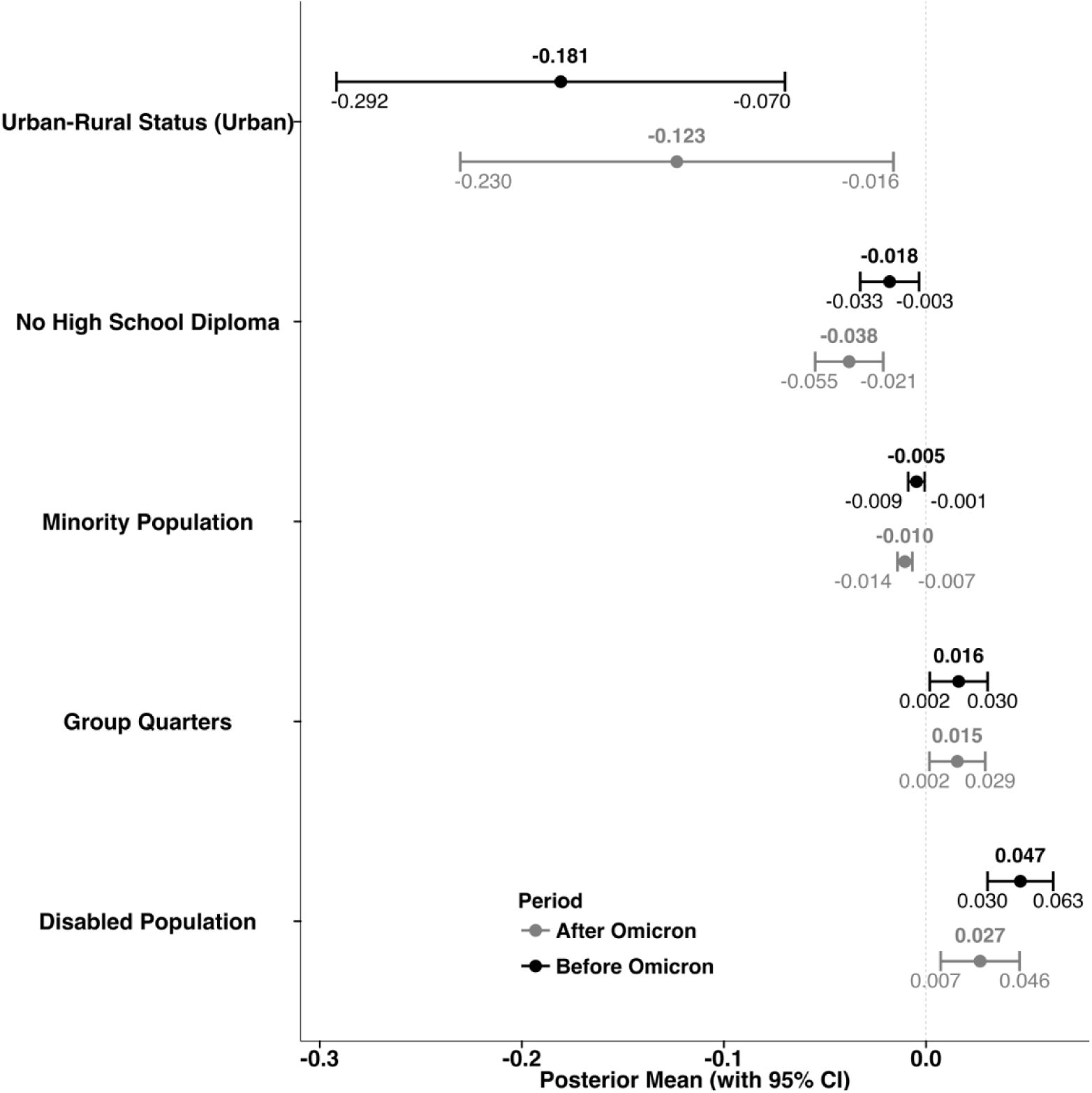
Posterior estimates (95% credible intervals (CIs)) of regression coefficients that measure the relationships between social vulnerability factors and long COVID incidence based on Bayesian spatial random effect models.

Similarly, counties with more residents living in group quarters showed a significant positive association in both periods (0.016, 0.002 to 0.030 before; 0.015, 0.002 to 0.029 after). Additional variables that were significantly associated with long COVID incidence but only in one period are shown in **Figure** S10. For example, the proportion of the population aged over 65 (–0.021, –0.034 to –0.008), multi-unit housing (–0.013, –0.022 to –0.005), limited English proficiency (0.040, 0.009 to 0.072), and the proportion below 150% of the federal poverty level (0.017, 0.006 to 0.027) were significant only after Omicron, whereas first vaccination rate (–0.006, –0.009 to –0.002) and housing cost burden (0.015, 0.003 to 0.028) were significant only before Omicron.

In particular, the spatial random effect methods improved model fit and strengthened the robustness of statistical inference based on histograms of residuals, along with the results of Anderson–Darling test (**Figure** S9 of the Supplementary Material).

### 3.5 Impacts of sociodemographic disparities on the long COVID incidence

Figure 4 (a-f) presents the temporal trends in long COVID incidence stratified by selected SVI-related factors, with patterns broadly consistent with the findings of spatial random effect models. Counties with a higher proportion of residents living below 150% of the poverty level (>12%) consistently exhibited higher long COVID incidence than those with lower poverty levels (≤12%) (Figure 4a). Similarly, counties with a greater proportion of disabled residents (>10%) experienced higher incidence compared to those with lower disability prevalence (≤10%) across most quarters (Figure 4b). In Figure 4c, counties with more residents living in group quarters (>3%) also showed elevated incidence relative to those with ≤3%. Interestingly, Figure 4d indicates that counties with higher minority population proportions (>20%) tended to have lower long COVID incidence than those with lower proportions (≤20%), particularly after 2022. Counties with higher vaccination rates (>60%) consistently reported lower incidence than those with lower rates (≤60%) (Figure 4e). Finally, rural counties exhibited higher long COVID incidence than urban counties throughout the study period, with a marked divergence following the emergence of Omicron (Figure 4f). In particular, the threshold for each variable was selected based on comparison analysis to ensure that, under the given threshold, the difference in long COVID incidence related to the variable was statistically significant in most of the six subregions (**Table** S6 of the Supplementary Material). This approach ensured the robustness of the results above.

**Figure 4.**
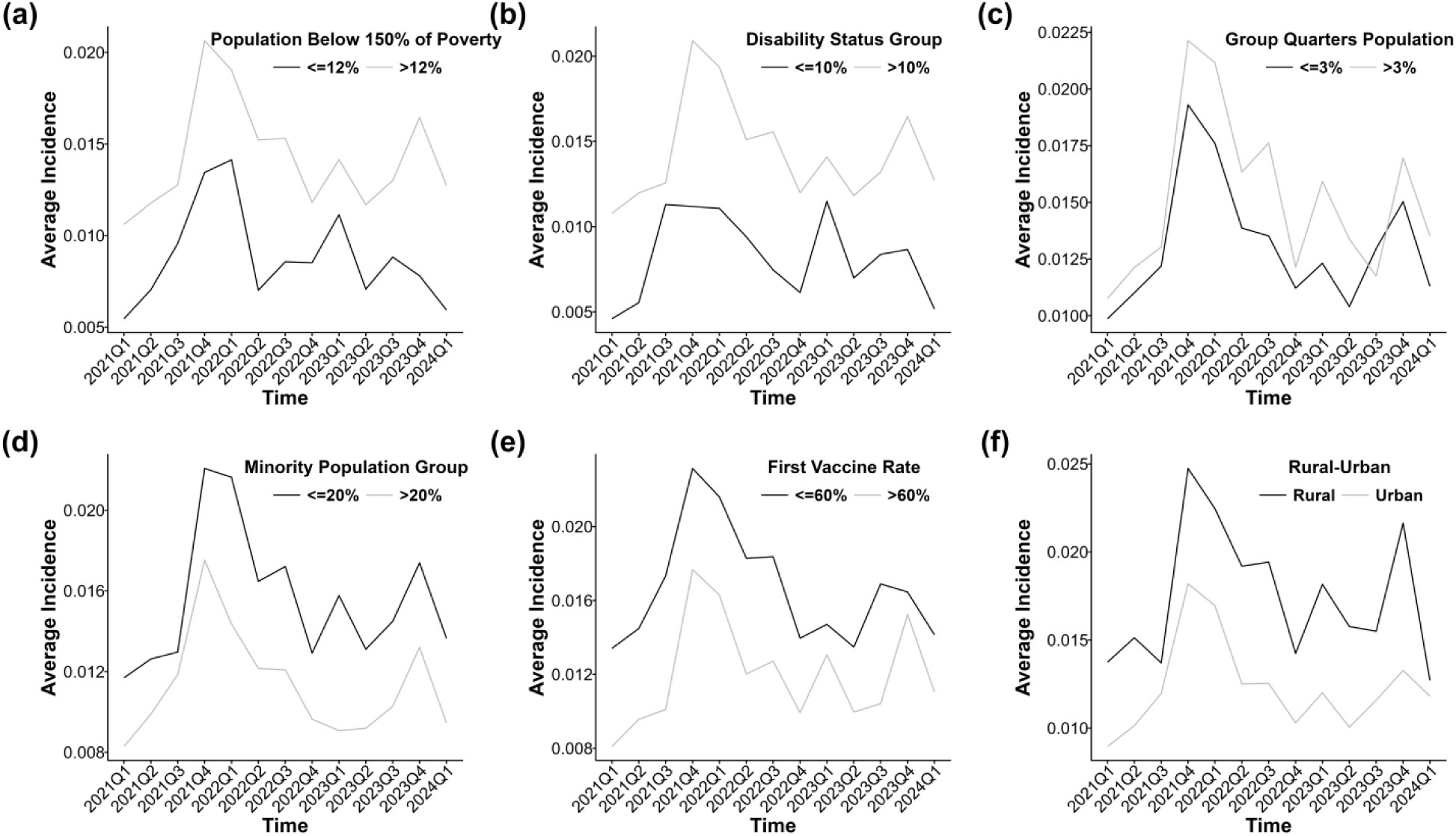
Temporal trends in long COVID incidence across different levels of six social vulnerability factors: (a) poverty status (population below 150% of the federal poverty level), (b) disability prevalence, (c) residence in group quarters, (d) racial and ethnic minority composition, (e) COVID-19 vaccination coverage, and (f) rural – urban classification.

## 4. Discussion

This study offers a comprehensive view of long COVID incidence by analyzing its spatiotemporal patterns and associations with area-level socioeconomic and demographic factors, extending beyond traditional individual-level analyses. Using a large U.S. dataset of 5,652,474 COVID-19 and 41,694 long COVID cases from 2021 to 2024, we identified significant spatiotemporal differences in its epidemiology. Spatial clustering of incidence was observed across many counties (p < 0.05). Notably, low-incidence areas were more concentrated on the U.S. East Coast, while high-incidence areas distributed inland regions, likely reflecting pre-existing disparities in health (healthcare and public health) services coverage and available social infrastructure. This epidemiological pattern may in part be explained by evidence of socioeconomic and demographic clustering in these regions, where Americans of higher socioeconomic status have been reported to be migrating to the coastal regions while those with lower socioeconomic status are moving to the inland regions^22^. These findings further suggest that low-income and underserved populations in inland regions may face a disproportionately higher risk of developing long COVID. This elevated vulnerability is likely driven by the higher prevalence of contributing factors in these communities, such as comorbidities and smoking^23,24^.

This study identifies SVI factors associated with long COVID and clarifies their role in spatiotemporal disparities. Our analysis identified rurality, poverty levels, disability prevalence, institutional living environments, minority composition, vaccination coverage, and residential crowding as key factors influencing long COVID incidence, with varying effects across time periods. Specifically, we consistently observed higher long COVID incidence between 2021 and 2024 in counties characterized by rural status, higher disability prevalence, and a larger share of residents in institutional living environments. Counties with higher poverty levels and greater residential crowding became more strongly associated with higher long COVID incidence after the emergence of the Omicron variant. This finding highlights the disproportionate burden of long COVID among economically vulnerable communities. Existing healthcare and social determinants of health inequities likely played a critical role in shaping these patterns, as lower healthcare access and higher economic vulnerability contribute to prolonged post-acute symptoms and delayed medical diagnosis and treatment^25^. In contrast, higher COVID-19 vaccination coverage was associated with lower long COVID incidence primarily in the pre-Omicron period, suggesting a potential early protective effect against long-term complications. Interestingly, counties with higher minority populations and lower educational attainment showed lower reported incidence. This may reflect underdiagnosis and ecological bias, as county-level metrics may not reflect the characteristics of individuals in the N3C dataset. Moreover, individuals from minority backgrounds or with lower education levels may be less likely to seek care for persistent post-COVID symptoms due to health literacy challenges, language barriers, limited healthcare access, or financial and structural obstacles. As a result, these populations are more likely to be underrepresented in EHR-based long COVID case definitions. As such, future studies leveraging N3C data should interpret these associations with caution and consider the potential impact of ecological bias and data representativeness when evaluating racial and ethnic disparities or levels of educational attainment in long COVID burden.

Given the significant spatiotemporal disparities in long COVID incidence, policy responses should promote equitable diagnosis and care. Targeted healthcare resource allocation with risk stratification is essential, with high-incidence areas requiring specialized long COVID clinics, rehabilitation centers, and increased funding for chronic disease prevention and control, while low-incidence areas should be assessed for under-reporting, under-diagnosis, clinician competency, and population awareness gaps^26^. Given that many high-risk regions are in underserved Midwestern areas, expanding telemedicine services and deploying mobile health units can improve accessibility to specialized care for acute COVID-19, long COVID, and existing comorbidities^27^. Additionally, in areas with limited healthcare facilities and low population engagement with the healthcare system for long COVID diagnosis and care, outreach programs and social support initiatives should be implemented to improve early detection and intervention and reduce the entrenched individual-level and social determinants of health inequities. Related policies, such as the disability measure^28^, can also be adjusted to reflect geographic and demographic disparities, ensuring adequate support for long COVID patients facing economic and healthcare barriers^26^. Addressing these disparities through policy-driven health interventions, including healthcare accessibility improvements and targeted public health interventions implementation will help mitigate the short-term and long-term impact of long COVID, ultimately leading to better health outcomes for vulnerable populations, reduced associated health costs for the governments, and improved health equity.

This study has several limitations. First, due to constraints in the N3C EHR database, approximately 35% of long COVID records lacked corresponding COVID-19 EHR records, likely because many patients experienced mild symptoms and did not seek healthcare, leading to a dataset biased toward individuals with more severe acute infections^29^. Second, consistent with data from most surveillance systems, the long COVID incidence estimated from N3C sentinel surveillance system might not represent the true burden of the disease due to under-ascertainment bias from symptomatic cases who do not seek health care due to healthcare access (physical and/or economic) constraints and poor awareness of long COVID symptoms. Third, sampling bias may have been introduced due to the limited number of sentinel sites included in the database, and another important limitation is the uneven distribution of these sites across U.S. regions, which may have led to some areas being oversampled while others were under-sampled. This imbalance could introduce regional biases in the data, potentially affecting observed incidence rates and risk factor associations, thereby reducing the generalizability of the findings to the broader U.S. population, particularly in regions with fewer or no sentinel sites where healthcare access and demographic characteristics may differ significantly. Fourth, our method for estimating incidence risks involved excluding patients who had not received a long COVID diagnosis within 180 days after their COVID-19 infection, which may have led to measurement bias resulting from an underestimation of the actual disease burden. Moreover, it is also possible that the differences in the data collection time period for the N3C EHR and SVI databases have contributed to measurement bias. Fifth, due to uneven data collection in the N3C EHR system, regions with well-resourced healthcare facilities may report higher long COVID diagnoses due to improved detection from robust screening and diagnostic practices, while under-resourced areas may underreport cases, potentially distorting geographic patterns. Sixth, to maintain data stability, we excluded counties with fewer than 20 COVID-19 records, which may have further affected spatial representation. Finally, since long COVID is an extremely heterogeneous condition encompassing a wide range of symptoms^30^, our study did not differentiate cases by specific symptom profiles, which may obscure important subgroup trends. Future research should explore symptom-specific patterns and pre-existing medical conditions to enhance the understanding of long COVID incidence and its spatiotemporal patterns and wider social determinants of health for improved design and delivery of tailored and targeted health and social interventions at both the population and healthcare system levels.

## Supplementary Material

Supplementary Material provided more details, including descriptions of the datasets, data processing, and additional results.

## Author Contributions

Z. Chen and B. Li had full access to all the data in the study and take responsibility for the integrity of the data and the accuracy of the data analysis.

*Concept and design:* Z. Chen, B. Li, Y. Chen, J. Liu, F. Luo, X. Chen, Y. Shen.

*Acquisition, analysis, or interpretation of data:* Z. Chen, B. Li, Y. Chen, J. Liu, F. Luo.

*Drafting of the manuscript:* Z. Chen, B. Li, Y. Chen, J. Liu, F. Luo.

*Critical revision of the manuscript for important intellectual content:* Z. Chen, B. Li, Y. Chen, J. Liu, F. Luo, K. Ogunyemi, Y. Ge, Y. Ke, Y. Yang, X. Chen, Y. Shen.

*Statistical analysis:* Z. Chen, B. Li, Y. Chen.

*Obtained funding:* Y. Shen.

*Administrative, technical, or material support:* X. Chen, Y. Shen.

*Supervision:* X. Chen, Y. Shen.

## Acknowledgments

The analyses described in this publication were conducted with data or tools accessed through the NCATS N3C Data Enclave https://covid.cd2h.org and N3C Attribution & Publication Policy v 1.2-2020-08-25b supported by NCATS Contract No. 75N95023D00001, Axle Informatics Subcontract: NCATS-P00438-B.

## Disclaimer

The N3C Publication committee confirmed that this manuscript MSID: 2441.041 is in accordance with N3C data use and attribution policies; however, this content is solely the responsibility of the authors and does not necessarily represent the official views of the National Institutes of Health or the N3C program.

## IRB

The IRB ID of this project is PROJECT00007079, and the Data Use Request (DUR) ID is DUR-6E0E8C9. The N3C Data Enclave is managed under the authority of the NIH; information can be found at https://ncats.nih.gov/n3c/resources.

## Individual Acknowledgements for Core Contributors

We gratefully acknowledge the following core contributors to N3C:

Adam B. Wilcox, Adam M. Lee, Alexis Graves, Alfred (Jerrod) Anzalone, Amin Manna, Amit Saha, Amy Olex, Andrea Zhou, Andrew E. Williams, Andrew M. Southerland, Andrew T. Girvin, Anita Walden, Anjali Sharathkumar, Benjamin Amor, Benjamin Bates, Brian Hendricks, Brijesh Patel, G. Caleb Alexander, Carolyn T. Bramante, Cavin Ward-Caviness, Charisse Madlock-Brown, Christine Suver, Christopher G. Chute, Christopher Dillon, Chunlei Wu, Clare Schmitt, Cliff Takemoto, Dan Housman, Davera Gabriel, David A. Eichmann, Diego Mazzotti, Donald E. Brown, Eilis Boudreau, Elaine L. Hill, Emily Carlson Marti, Emily R. Pfaff, Evan French, Farrukh M Koraishy, Federico Mariona, Fred Prior, George Sokos, Greg Martin, Harold P. Lehmann, Heidi Spratt, Hemalkumar B. Mehta, J.W. Awori Hayanga, Jami Pincavitch, Jaylyn Clark, Jeremy Richard Harper, Jessica Yasmine Islam, Jin Ge, Joel Gagnier, Johanna J. Loomba, John B. Buse, Jomol Mathew, Joni L. Rutter, Julie A. McMurry, Justin Guinney, Justin Starren, Karen Crowley, Katie Rebecca Bradwell, Kellie M. Walters, Ken Wilkins, Kenneth R. Gersing, Kenrick Cato, Kimberly Murray, Kristin Kostka, Lavance Northington, Lee Pyles, Lesley Cottrell, Lili M. Portilla, Mariam Deacy, Mark M. Bissell, Marshall Clark, Mary Emmett, Matvey B. Palchuk, Melissa A. Haendel, Meredith Adams, Meredith Temple-O’Connor, Michael G. Kurilla, Michele Morris, Nasia Safdar, Nicole Garbarini, Noha Sharafeldin, Ofer Sadan, Patricia A. Francis, Penny Wung Burgoon, Philip R.O. Payne, Randeep Jawa, Rebecca Erwin-Cohen, Rena C. Patel, Richard A. Moffitt, Richard L. Zhu, Rishikesan Kamaleswaran, Robert Hurley, Robert T. Miller, Saiju Pyarajan, Sam G. Michael, Samuel Bozzette, Sandeep K. Mallipattu, Satyanarayana Vedula, Scott Chapman, Shawn T. O’Neil, Soko Setoguchi, Stephanie S. Hong, Steven G. Johnson, Tellen D. Bennett, Tiffany J. Callahan, Umit Topaloglu, Valery Gordon, Vignesh Subbian, Warren A. Kibbe, Wenndy Hernandez, Will Beasley, Will Cooper, William Hillegass, Xiaohan Tanner Zhang. Details of contributions available at https://covid.cd2h.org/core-contributors

## Data Partners with Released Data

The following institutions whose data is released or pending:

Available: Advocate Health Care Network — UL1TR002389: The Institute for Translational Medicine (ITM) • Aurora Health Care Inc — UL1TR002373: Wisconsin Network For Health Research • Boston University Medical Campus — UL1TR001430: Boston University Clinical and Translational Science Institute • Brown University — U54GM115677: Advance Clinical Translational Research (Advance-CTR) • Carilion Clinic — UL1TR003015: iTHRIV Integrated Translational health Research Institute of Virginia • Case Western Reserve University — UL1TR002548: The Clinical& Translational Science Collaborative of Cleveland (CTSC) • Charleston Area Medical Center — U54GM104942: West Virginia Clinical and Translational Science Institute (WVCTSI) • Children’s Hospital Colorado — UL1TR002535: Colorado Clinical and Translational Sciences Institute • Columbia University Irving Medical Center — UL1TR001873: Irving Institute for Clinical and Translational Research • Dartmouth College — None (Voluntary) Duke University — UL1TR002553: Duke Clinical and Translational Science Institute • George Washington Children’s Research Institute — UL1TR001876: Clinical and Translational Science Institute at Children’s National (CTSA-CN) • George Washington University — UL1TR001876: Clinical and Translational Science Institute at Children’s National (CTSA-CN) • Harvard Medical School — UL1TR002541: Harvard Catalyst • Indiana University School of Medicine — UL1TR002529: Indiana Clinical and Translational Science Institute • Johns Hopkins University — UL1TR003098: Johns Hopkins Institute for Clinical and Translational Research • Louisiana Public Health Institute — None (Voluntary) • Loyola Medicine — Loyola University Medical Center • Loyola University Medical Center — UL1TR002389: The Institute for Translational Medicine (ITM) • Maine Medical Center — U54GM115516: Northern New England Clinical & Translational Research (NNE-CTR) Network • Mary Hitchcock Memorial Hospital & Dartmouth Hitchcock Clinic — None (Voluntary) • Massachusetts General Brigham — UL1TR002541: Harvard Catalyst • Mayo Clinic Rochester — UL1TR002377: Mayo Clinic Center for Clinical and Translational Science (CCaTS) • Medical University of South Carolina — UL1TR001450: South Carolina Clinical & Translational Research Institute (SCTR) • MITRE Corporation — None (Voluntary) • Montefiore Medical Center — UL1TR002556: Institute for Clinical and Translational Research at Einstein and Montefiore • Nemours — U54GM104941: Delaware CTR ACCEL Program • NorthShore University HealthSystem — UL1TR002389: The Institute for Translational Medicine (ITM) • Northwestern University at Chicago — UL1TR001422: Northwestern University Clinical and Translational Science Institute (NUCATS) • OCHIN — INV-018455: Bill and Melinda Gates Foundation grant to Sage Bionetworks • Oregon Health & Science University — UL1TR002369: Oregon Clinical and Translational Research Institute • Penn State Health Milton S. Hershey Medical Center — UL1TR002014: Penn State Clinical and Translational Science Institute • Rush University Medical Center — UL1TR002389: The Institute for Translational Medicine (ITM) • Rutgers, The State University of New Jersey — UL1TR003017: New Jersey Alliance for Clinical and Translational Science • Stony Brook University — U24TR002306 • The Alliance at the University of Puerto Rico, Medical Sciences Campus — U54GM133807: Hispanic Alliance for Clinical and Translational Research (The Alliance) • The Ohio State University — UL1TR002733: Center for Clinical and Translational Science • The State University of New York at Buffalo — UL1TR001412: Clinical and Translational Science Institute • The University of Chicago — UL1TR002389: The Institute for Translational Medicine (ITM) • The University of Iowa — UL1TR002537: Institute for Clinical and Translational Science • The University of Miami Leonard M. Miller School of Medicine — UL1TR002736: University of Miami Clinical and Translational Science Institute • The University of Michigan at Ann Arbor — UL1TR002240: Michigan Institute for Clinical and Health Research • The University of Texas Health Science Center at Houston — UL1TR003167: Center for Clinical and Translational Sciences (CCTS) • The University of Texas Medical Branch at Galveston — UL1TR001439: The Institute for Translational Sciences • The University of Utah — UL1TR002538: Uhealth Center for Clinical and Translational Science • Tufts Medical Center — UL1TR002544: Tufts Clinical and Translational Science Institute • Tulane University — UL1TR003096: Center for Clinical and Translational Science • The Queens Medical Center — None (Voluntary) • University Medical Center New Orleans — U54GM104940: Louisiana Clinical and Translational Science (LA CaTS) Center • University of Alabama at Birmingham — UL1TR003096: Center for Clinical and Translational Science • University of Arkansas for Medical Sciences — UL1TR003107: UAMS Translational Research Institute • University of Cincinnati — UL1TR001425: Center for Clinical and Translational Science and Training • University of Colorado Denver, Anschutz Medical Campus — UL1TR002535: Colorado Clinical and Translational Sciences Institute • University of Illinois at Chicago — UL1TR002003: UIC Center for Clinical and Translational Science • University of Kansas Medical Center — UL1TR002366: Frontiers: University of Kansas Clinical and Translational Science Institute • University of Kentucky — UL1TR001998: UK Center for Clinical and Translational Science • University of Massachusetts Medical School Worcester — UL1TR001453: The UMass Center for Clinical and Translational Science (UMCCTS) • University Medical Center of Southern Nevada — None (voluntary) • University of Minnesota — UL1TR002494: Clinical and Translational Science Institute • University of Mississippi Medical Center — U54GM115428: Mississippi Center for Clinical and Translational Research (CCTR) • University of Nebraska Medical Center — U54GM115458: Great Plains IDeA-Clinical & Translational Research • University of North Carolina at Chapel Hill — UL1TR002489: North Carolina Translational and Clinical Science Institute • University of Oklahoma Health Sciences Center — U54GM104938: Oklahoma Clinical and Translational Science Institute (OCTSI) • University of Pittsburgh — UL1TR001857: The Clinical and Translational Science Institute (CTSI) • University of Pennsylvania — UL1TR001878: Institute for Translational Medicine and Therapeutics • University of Rochester — UL1TR002001: UR Clinical & Translational Science Institute • University of Southern California — UL1TR001855: The Southern California Clinical and Translational Science Institute (SC CTSI) • University of Vermont — U54GM115516: Northern New England Clinical & Translational Research (NNE-CTR) Network • University of Virginia — UL1TR003015: iTHRIV Integrated Translational health Research Institute of Virginia • University of Washington — UL1TR002319: Institute of Translational Health Sciences • University of Wisconsin-Madison — UL1TR002373: UW Institute for Clinical and Translational Research • Vanderbilt University Medical Center — UL1TR002243: Vanderbilt Institute for Clinical and Translational Research • Virginia Commonwealth University — UL1TR002649: C. Kenneth and Dianne Wright Center for Clinical and Translational Research • Wake Forest University Health Sciences — UL1TR001420: Wake Forest Clinical and Translational Science Institute • Washington University in St. Louis — UL1TR002345: Institute of Clinical and Translational Sciences • Weill Medical College of Cornell University — UL1TR002384: Weill Cornell Medicine Clinical and Translational Science Center • West Virginia University — U54GM104942: West Virginia Clinical and Translational Science Institute (WVCTSI) • Submitted: Icahn School of Medicine at Mount Sinai — UL1TR001433: ConduITS Institute for Translational Sciences • The University of Texas Health Science Center at Tyler — UL1TR003167: Center for Clinical and Translational Sciences (CCTS) • University of California, Davis — UL1TR001860: UCDavis Health Clinical and Translational Science Center • University of California, Irvine — UL1TR001414: The UC Irvine Institute for Clinical and Translational Science (ICTS) • University of California, Los Angeles — UL1TR001881: UCLA Clinical Translational Science Institute • University of California, San Diego — UL1TR001442: Altman Clinical and Translational Research Institute • University of California, San Francisco — UL1TR001872: UCSF Clinical and Translational Science Institute NYU Langone Health Clinical Science Core, Data Resource Core, and PASC Biorepository Core — OTA-21-015A: Post-Acute Sequelae of SARS-CoV-2 Infection Initiative (RECOVER) Pending: Arkansas Children’s Hospital — UL1TR003107: UAMS Translational Research Institute • Baylor College of Medicine — None (Voluntary) • Children’s Hospital of Philadelphia — UL1TR001878: Institute for Translational Medicine and Therapeutics • Cincinnati Children’s Hospital Medical Center — UL1TR001425: Center for Clinical and Translational Science and Training • Emory University — UL1TR002378: Georgia Clinical and Translational Science Alliance • HonorHealth — None (Voluntary) • Loyola University Chicago — UL1TR002389: The Institute for Translational Medicine (ITM) • Medical College of Wisconsin — UL1TR001436: Clinical and Translational Science Institute of Southeast Wisconsin • MedStar Health Research Institute — None (Voluntary) • Georgetown University — UL1TR001409: The Georgetown-Howard Universities Center for Clinical and Translational Science (GHUCCTS) • MetroHealth — None (Voluntary) • Montana State University — U54GM115371: American Indian/Alaska Native CTR • NYU Langone Medical Center — UL1TR001445: Langone Health’s Clinical and Translational Science Institute • Ochsner Medical Center — U54GM104940: Louisiana Clinical and Translational Science (LA CaTS) Center • Regenstrief Institute — UL1TR002529: Indiana Clinical and Translational Science Institute • Sanford Research — None (Voluntary) • Stanford University — UL1TR003142: Spectrum: The Stanford Center for Clinical and Translational Research and Education • The Rockefeller University — UL1TR001866: Center for Clinical and Translational Science • The Scripps Research Institute — UL1TR002550: Scripps Research Translational Institute • University of Florida — UL1TR001427: UF Clinical and Translational Science Institute — University of New Mexico Health Sciences Center — UL1TR001449: University of New Mexico Clinical and Translational Science Center • University of Texas Health Science Center at San Antonio — UL1TR002645: Institute for Integration of Medicine and Science • Yale New Haven Hospital — UL1TR001863: Yale Center for Clinical Investigation.

## Data availability

All data used in this study is available through the N3C Enclave to approved users. See https://covid.cd2h.org/for-researchers for instructions on how to access the data. We used N3C data from version 185.

## Supplementary Material

## S1 EHR of long COVID and data processing

Given the inherent characteristics of electronic health record (EHR) data, which primarily capture individuals with more severe symptoms who seek in-person medical care, our long COVID cohort is likely biased toward patients experiencing more severe or persistent symptoms. Notably, approximately 40% of identified long COVID cases lack corresponding acute COVID-19 records in the EHR, likely due to patients experiencing mild or asymptomatic acute infections and recovering at home without medical intervention, resulting in an absence of documented acute disease. As our objective is to estimate and compare the incidence risk of long COVID across regions, we define the at-risk population as individuals with a confirmed acute COVID-19 diagnosis, allowing for standardized denominator calculations. long COVID cases without a documented acute infection are therefore excluded to ensure the validity of risk estimation. This exclusion is essential for minimizing misclassification bias, as the absence of an acute COVID-19 record precludes accurate determination of symptom onset relative to infection, leading to potential inflation or deflation of risk estimates. Additionally, these cases may represent individuals who sought care only for persistent symptoms rather than those with a well-defined transition from acute infection to long COVID, introducing heterogeneity that could bias comparisons across regions. Furthermore, the completeness of acute COVID-19 documentation varies across healthcare systems, with differences in testing accessibility and healthcare-seeking behaviors influencing EHR records. By restricting the cohort to individuals with documented acute infections, we enhance the comparability of risk estimates across populations and mitigate potential confounding arising from regional variations in EHR capture. This approach ensures a robust and interpretable assessment of long COVID incidence following acute infection.

For identifying long COVID cases, we utilized ICD-10 codes B94.8 and U09.9 as diagnostic indicators. Since U09.9 was officially introduced as the long COVID diagnosis code on October 1, 2021, diagnoses related to long COVID prior to this date predominantly used the B94.8 code. Therefore, we applied a temporal cutoff on October 1, 2021, selecting EHR records with B94.8 diagnoses before this date and U09.9 diagnoses thereafter. This approach ensures comprehensive coverage of long COVID cases across different time periods, accounting for the evolution of diagnostic practices.

We initiated data cleaning using the original N3C dataset. Overall cleaning process was summarized in **Figure S1**. On one hand, COVID-19 patient records were cleaned, and on the other, long COVID patient records were similarly processed. Only records corresponding to a patient’s first COVID-19 infection or first long COVID infection were retained. In the process of filtering N3C EHR data, we also applied site-level selection criteria. During data cleaning, we observed that certain sites had ceased their collaboration with N3C by the time long COVID cases began to emerge. Including EHR data from these sites could potentially bias our analysis by underestimating the incidence of long COVID, thereby affecting subsequent data interpretation. To ensure data quality and consistency, we excluded sites with a long COVID incidence rate below 0.1%, as we deemed these sites invalid for analysis due to insufficient case reporting or incomplete data coverage. This site-level filtering helps improve the robustness of our incidence estimates and ensures more reliable regional comparisons.

**Figure S1:**
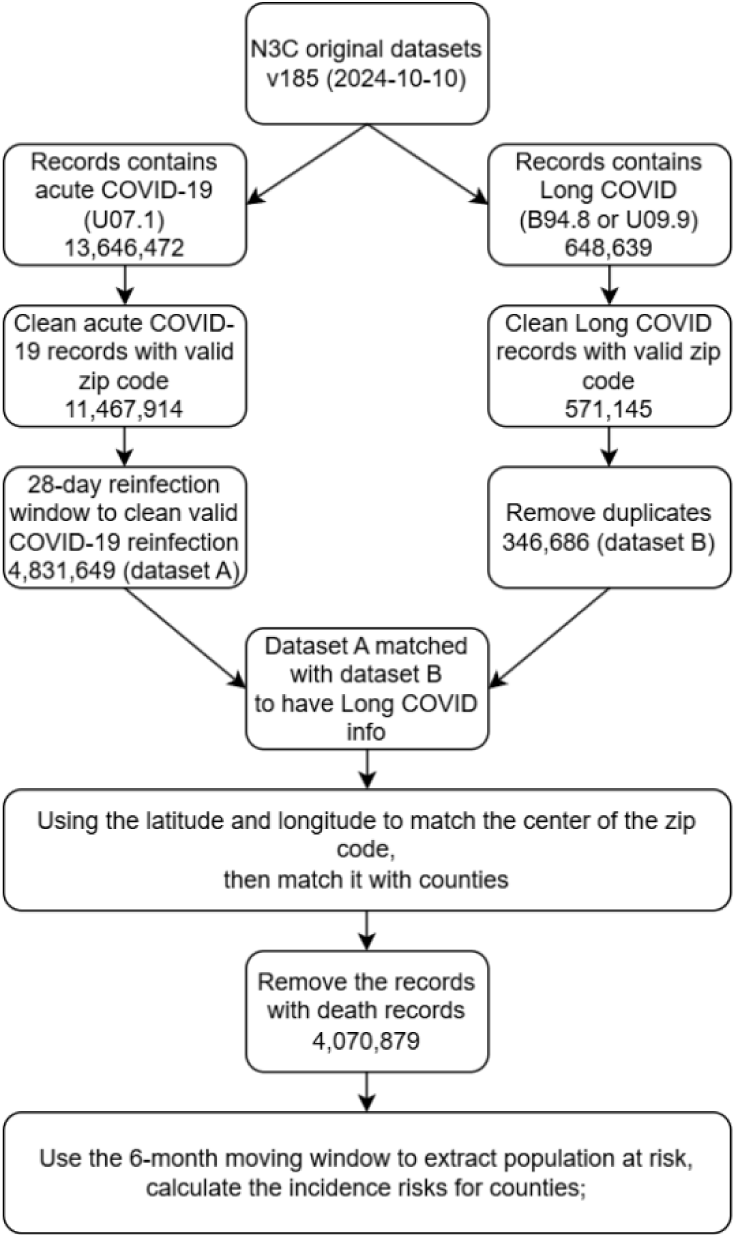
Diagram of data filtering and cleaning.

## S2 Dynamic incidence risk and definition of long COVID incidence

It is important to emphasize that all long COVID incidence calculations in this study are based exclusively on individuals who were previously diagnosed with acute COVID-19. That is, the at-risk population for all incidence metrics consistently includes only those with documented acute COVID-19. For the dynamic incidence risk algorithm shown in **Figure S1**, the criteria for defining the dynamic population at risk are as follows:

(a) **Earliest inclusion date for the population at risk:** We determined the start of the population at risk as 180 days prior to the earliest date of the target period. For example, for long COVID cases identified between January 1, 2021, and March 31, 2021, the corresponding population at risk consists of COVID-19 patients recorded in the EHR starting from July 1, 2020.
(b) **Latest inclusion date for the population at risk:** Following CDC guidelines, we adopted a one-month timeframe as the allowable window between the COVID-19 diagnosis and the long COVID diagnosis. The study [Crowd-sourced machine learning prediction of long COVID using data from the National COVID Cohort Collaborative] suggests that even when using B94.8 and U09.9 codes, a more conservative approach can be applied by using a 28-day threshold as a filter to refine long COVID data.

**Figure S2:**
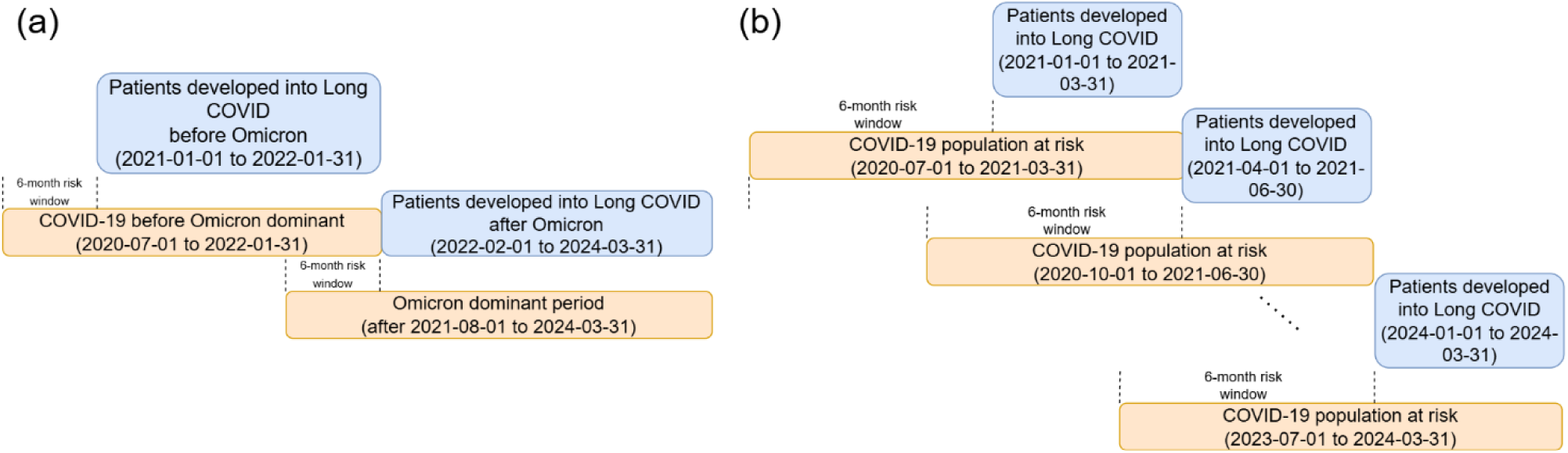
Dynamic incidence risk calculation. (a) The incidence of long COVID before and after the emergence of the Omicron variant in January 2022; and (b) The quarterly incidence of long COVID between 2021 and 2024.

In calculating these outcomes, we set a threshold of 180 days as a risk window to calculate dynamic incidence risk. This choice was made based on our observations from the N3C long COVID EHR and previous findings^1^, where most long COVID cases (over 77%) following an acute COVID-19 infection were diagnosed within 180 days of post-infection (Figure S3 of the Supplementary Material). If a patient was not diagnosed with long COVID within this threshold, we considered them no longer at risk of developing long COVID. Based on the guidance on t e International Classification of Diseases-10th Revision-Clinical Modification (ICD-10-CM), we identified acute COVID-19 using the U07.1 code to match our long-COVID definition, which relies on the B94.8 or U09.9 codes because long COVID has no corresponding biomarker tests^2^. To ensure a robust estimation of long COVID incidence during the dynamic incidence risk calculation, each time interval of interest was required to include at least 20 COVID-19 cases^3^.

**Figure S3:**
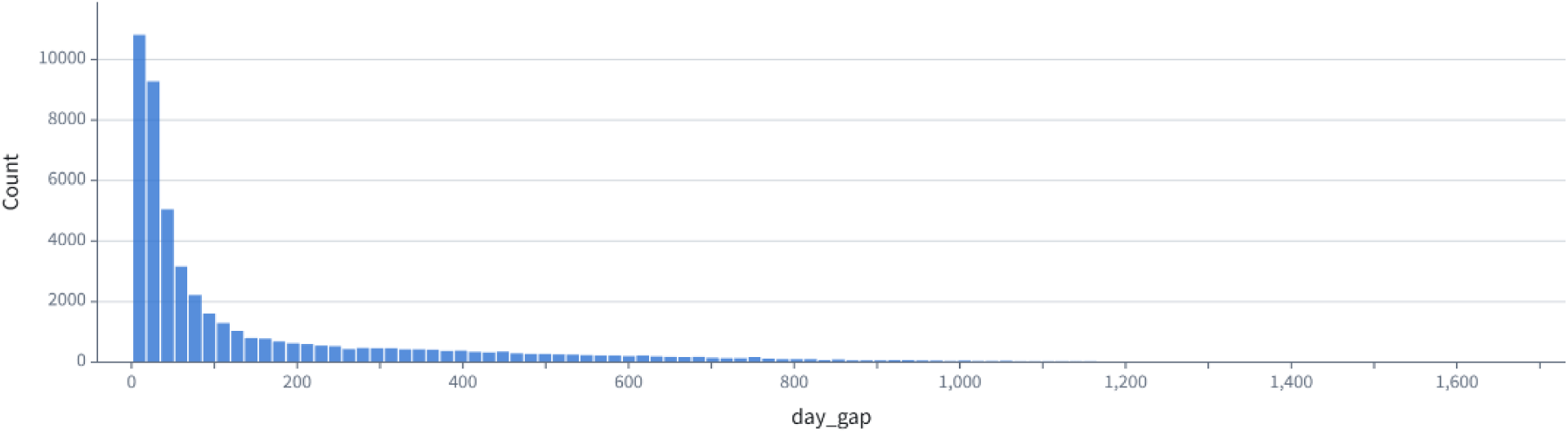
Summary histogram of gap days between acute COVID-19 diagnosis and long COVID diagnosis

## S3 Data exploration analysis

For each county, the local Moran’s I was computed as:

where     is the total number of U.S. counties,     is the incidence in county, ---- is the average incidence across counties,     is the variance of incidences, and     is the spatial weight between counties      and     . The weights were defined based on semivariogram analysis results using an exponential function (**Figure** S4). Statistical significance for both methods was assessed using a Monte Carlo simulation by generating a pseudo-random distribution to determine whether the observed Moran’s I and Getis is significantly different from randomness^4^.

**Figure S4:**
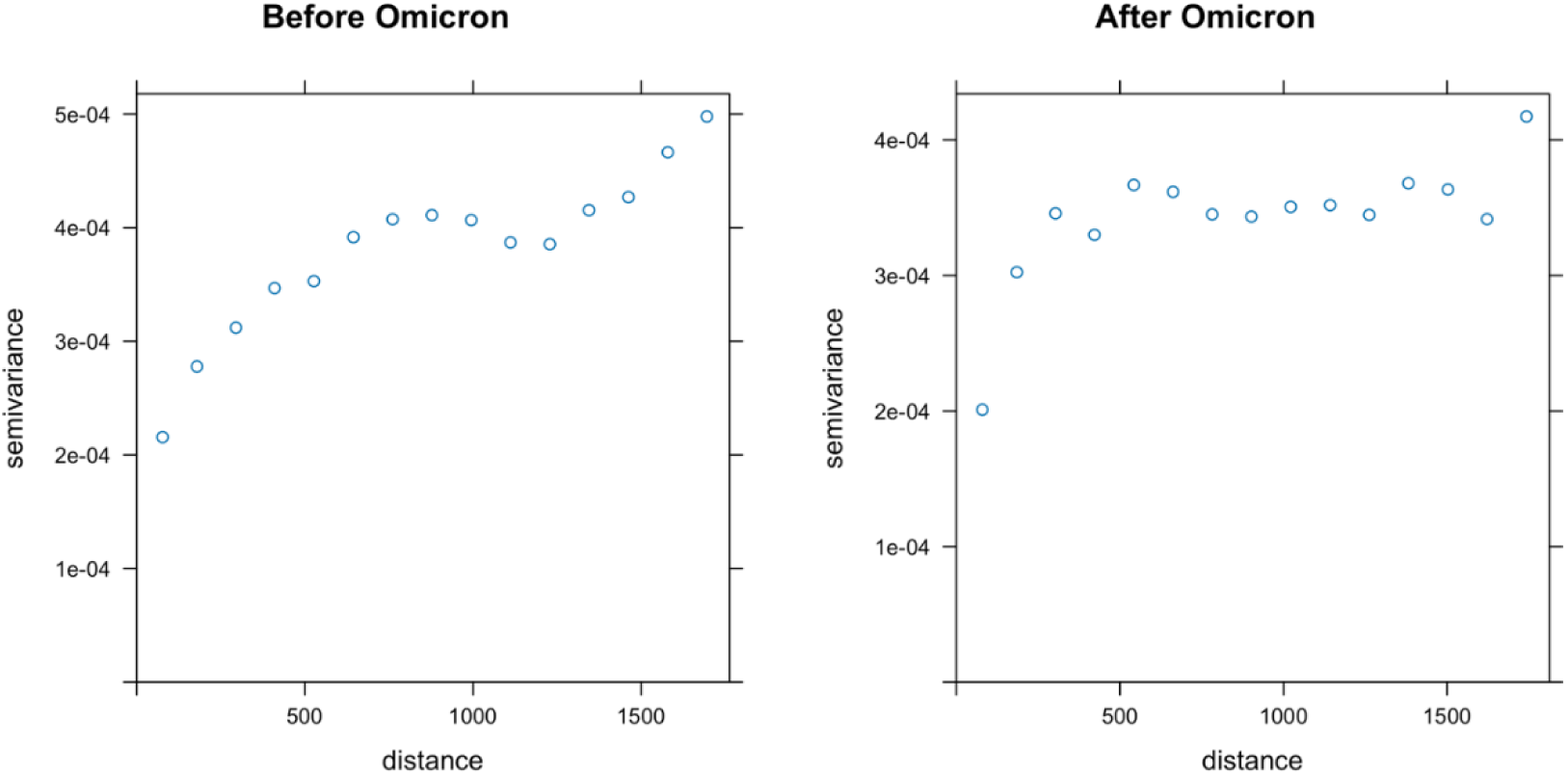
Semi-variogram analysis of county mean incidence before and after Omicron

**Figure S5:**
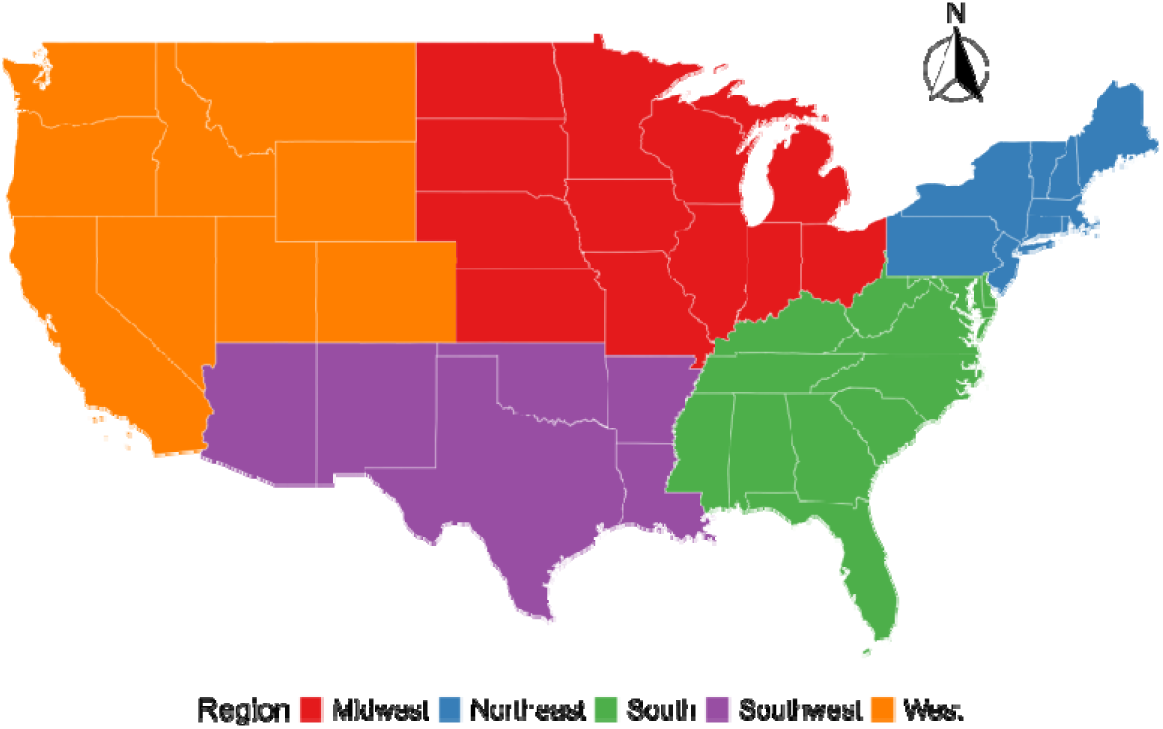
Five-subregion division of the United States

## S4 Long COVID incidence patterns incidence before and after Omicron dominance across different subregions of the United States

**Table S1.** Changes in the number of counties with long COVID incidence before and after Omicron dominance across different subregions of the United States (n = 673 counties).

| Region | The number of counties |  |  |  |
| --- | --- | --- | --- | --- |
|  | Decreased | Increased | No Change | Increased Rate<sup>a</sup> (%) |
| Northeast | 15 | 45 | 0 | 75.00 |
| West | 27 | 63 | 0 | 70.00 |
| Midwest | 72 | 165 | 0 | 69.62 |
| Southwest | 11 | 23 | 0 | 67.65 |
| South | 83 | 168 | 1 | 66.67 |
| Total | 208 | 464 | 1 | 68.95 |

<sup>a</sup>Increased Rate = Increased / (Increased + Decreased + No Change) \* 100.

**Table S2:** Average of long COVID incidence before and after Omicron across different subregions of the United States.

| Region | Average incidence across counties |  |
| --- | --- | --- |
|  | Before (%) | After (%) |
| Northeast | 1.66 | 2.45 |
| West | 2.42 | 2.86 |
| Midwest | 2.23 | 2.79 |
| Southwest | 1.85 | 1.78 |
| South | 2.21 | 2.53 |

**Figure S6:**
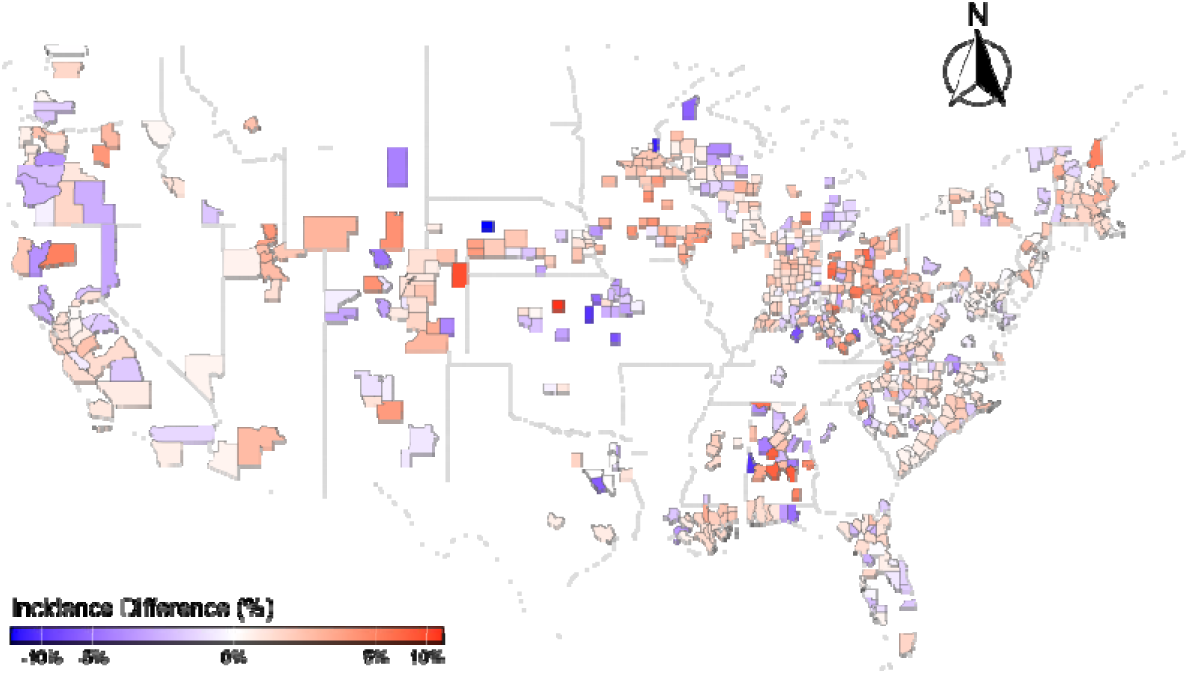
The map for the difference in average incidences between before and after t e dominance of Omicron (i.e., January 2022).

**Table S3.** Regional comparison of significant local Moran’s I before and after Omicron (n = 673 counties).

| Region | The number (rate<sup>a</sup>) of counties with positive correlation |  | The number (rate) of counties with negative correlation |  |
| --- | --- | --- | --- | --- |
|  | Before | After | Before | After |
| South | 130 (51.6%) | 128 (50.8%) | 37 (14.7%) | 36 (14.3%) |
| Northeast | 26 (43.3%) | 13 (21.7%) | 5 (8.3%) | 3 (5.0%) |
| Midwest | 54 (22.8%) | 55 (23.2%) | 17 (7.2%) | 30 (12.7%) |
| West | 10 (11.1%) | 29 (32.2%) | 10 (11.1%) | 18 (20.0%) |
| Southwest | 0 (0.0%) | 13 (38.2%) | 4 (11.8%) | 3 (8.8%) |
| Total | 220 (32.7%) | 238 (35.4%) | 73 (10.8%) | 90 (13.4%) |

<sup>a</sup> Rate = (Number of significant counties) / (Total number of counties in the region).

**Figure S7:**
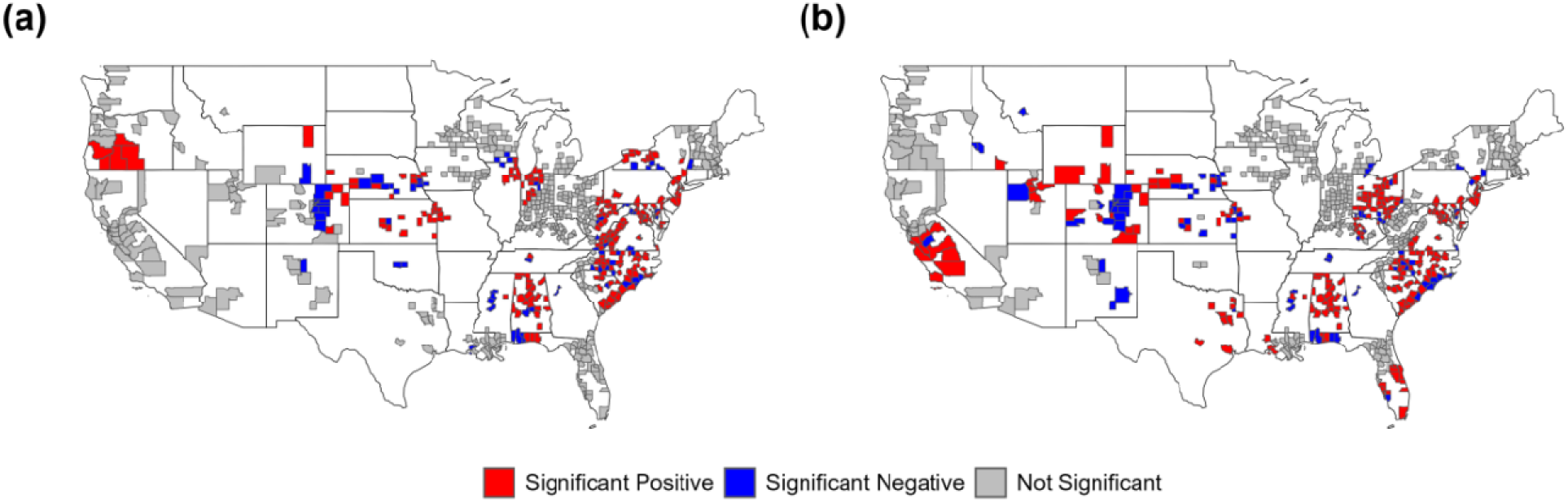
Spatial correlation of long COVID incidence across U.S. counties assessed using local Moran’s I: (a) before and (b) after Omicron dominance.

## S5 Stepwise Regression

We first adopt stepwise regression model to identify a subset of variables that best explain the variation in our study outcome, i.e., incidence of long COVID. To stable variance of incidence, we employed a logarithmic transformation for incidence (Figure S8). The results from stepwise regression were present to the following Tables S4-S5.

**Figure S8:**
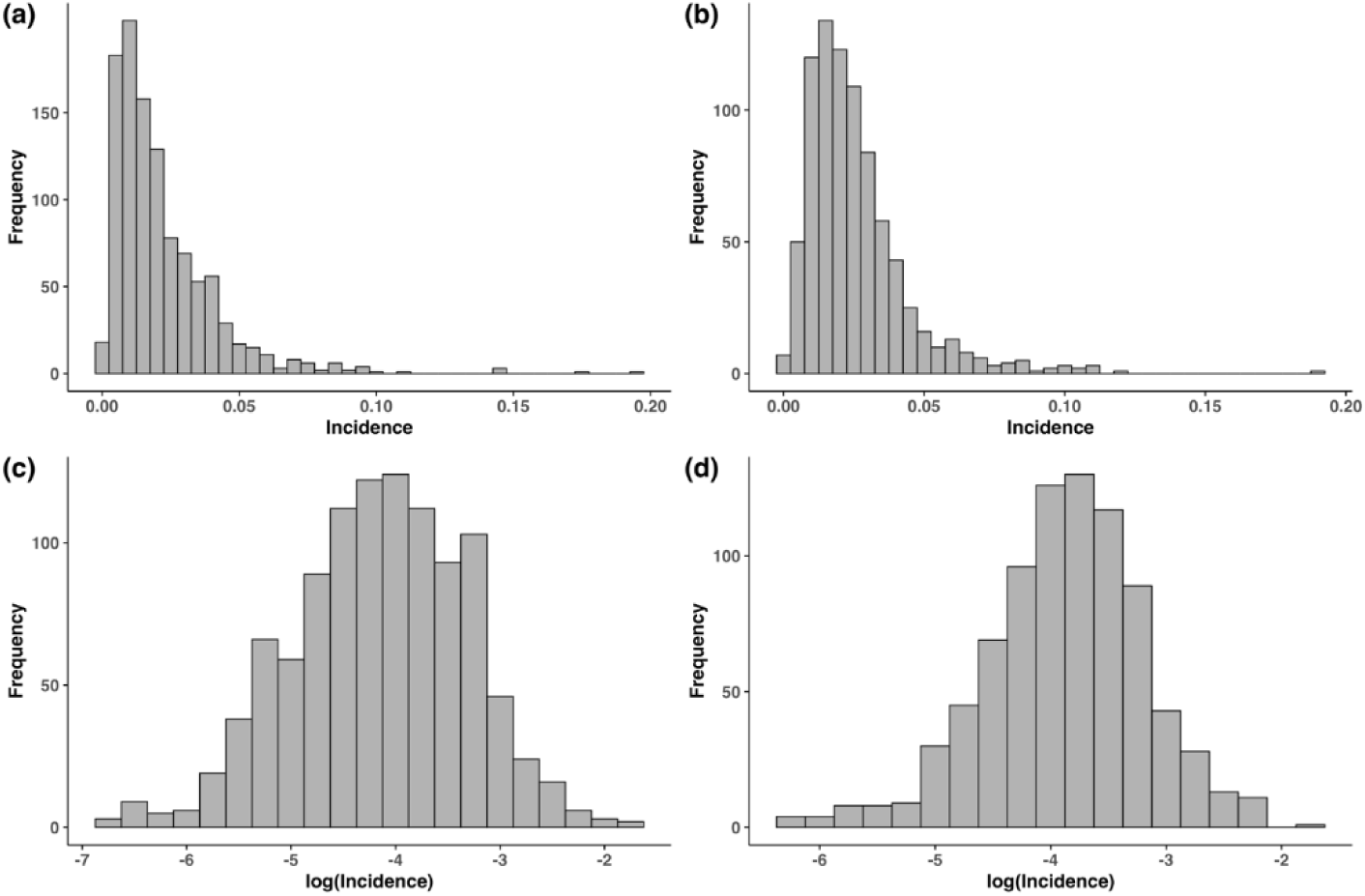
Distribution of county-level long COVID incidence before (a) and after (b) Omicron dominance, and the corresponding log-transformed counterparts, (c) and (d).

**Table S4.** Stepwise regression results for long COVID incidence before Omicron dominance.

| Estimate<br>(95% CI)<br>p-value | Model 1 | Model 2 | Model 3 | Model 4 | Model 5 |
| --- | --- | --- | --- | --- | --- |
| Disabled<br>Population<sup>a</sup> | 0.046<br>(0.035, 0.058)<br><.001* | 0.037<br>(0.024, 0.049)<br><.001* | 0.036<br>(0.023, 0.049)<br><.001* | 0.047<br>(0.031, 0.062)<br><.001* | 0.045<br>(0.026, 0.064)<br><.001* |
| Minority<br>Population<sup>b</sup> | -0.006<br>(-0.009, -0.004)<br><.001* | -0.008<br>(-0.011, -0.005)<br><.001* | -0.007<br>(-0.010, -0.004)<br><.001* | -0.004<br>(-0.008, -0.001)<br>0.024* | -0.005<br>(-0.009, -0.001)<br>0.008* |
| Urban-Rural<br>Status (Urban) | — | -0.222<br>(-0.331, -0.114)<br><.001* | -0.201<br>(-0.311, -0.091)<br><.001* | -0.203<br>(-0.315, -0.091)<br><.001* | -0.193<br>(-0.305, -0.080)<br><.001* |
| Housing Cost<br>Burden | — | 0.017<br>(0.005, 0.028)<br>0.004* | 0.024<br>(0.012, 0.036)<br><.001* | 0.020<br>(0.008, 0.032)<br>0.001* | 0.020<br>(0.008, 0.033)<br>0.002* |
| No Vehicle<br>Available | — | — | -0.017<br>(-0.028, -0.005)<br>0.005* | -0.015<br>(-0.027, -0.003)<br>0.013* | -0.013<br>(-0.025, -0.001)<br>0.029* |
| First Vaccination<br>Rate | — | — | -0.004<br>(-0.008, -0.001)<br>0.016* | -0.005<br>(-0.009, -0.001)<br>0.007* | -0.006<br>(-0.009, -0.002)<br>0.004* |
| No High School<br>Diploma | — | — | — | -0.018<br>(-0.032, -0.003)<br>0.015* | -0.029<br>(-0.046, -0.011)<br>0.001* |
| Group Quarters | — | — | — | 0.015<br>(0.001, 0.030)<br>0.036* | 0.015<br>(0.000, 0.030)<br>0.044* |
| Crowded Housing | — | — | — | — | 0.033<br>(-0.008, 0.075)<br>0.117 |
| Mobile Homes | — | — | — | — | 0.006<br>(-0.002, 0.014)<br>0.122 |

<sup>a</sup> Disability refers to the proportion of the civilian noninstitutionalized population with any disability; <sup>b</sup> The minority status refers to the percentage of the population that is not non-Hispanic White.

**Table S5.** Stepwise regression results for long COVID incidence after Omicron dominance.

| OR<br>(95%CI)<br>p-value | Model 1 | Model 2 | Model 3 | Model 4 | Model 5 |
| --- | --- | --- | --- | --- | --- |
| Minority<br>Population | -0.012<br>(-0.015, -0.010)<br><.001* | -0.010<br>(-0.012, -0.007)<br><.001* | -0.009<br>(-0.012, -0.006)<br><.001* | -0.009<br>(-0.013, -0.006)<br><.001* | -0.010<br>(-0.013, -0.006)<br><.001* |
| Below 150%<br>Poverty<br>Level | 0.015<br>(0.009, 0.021)<br><.001* | 0.018<br>(0.010, 0.027)<br><.001* | 0.012<br>(0.002, 0.023)<br>0.020* | 0.013<br>(0.002, 0.023)<br>0.019* | 0.014<br>(0.003, 0.025)<br>0.011* |
| Urban-Rural<br>Status<br>(Urban) | — | -0.160<br>(-0.264, -0.056)<br>0.003* | -0.187<br>(-0.293, -0.080)<br><.001* | -0.173<br>(-0.280, -0.067)<br>0.001* | -0.141<br>(-0.249, -0.033)<br>0.010* |
| No High<br>School<br>Diploma | — | -0.019<br>(-0.032, -0.006)<br>0.003* | -0.023<br>(-0.036, -0.010)<br><.001* | -0.039<br>(-0.055, -0.023)<br><.001* | -0.047<br>(-0.064, -0.030)<br><.001* |
| Population<br>Age 65+ | — | — | -0.017<br>(-0.030, -0.005)<br>0.008* | -0.017<br>(-0.029, -0.004)<br>0.011* | -0.019<br>(-0.032, -0.006)<br>0.005* |
| Disabled<br>Population<sup>b</sup> | — | — | 0.021<br>(0.003, 0.040)<br>0.023* | 0.031<br>(0.012, 0.051)<br>0.002* | 0.031<br>(0.011, 0.050)<br>0.002* |
| Limited<br>English | — | — | — | 0.039<br>(0.011, 0.068)<br>0.007* | 0.056<br>(0.026, 0.087)<br><.001* |
| First<br>Vaccination<br>Rate | — | — | — | -0.005<br>(-0.009, -0.001)<br>0.010* | -0.004<br>(-0.008, 0.000)<br>0.052 |
| Group<br>Quarters | — | — | — | — | 0.016<br>(0.002, 0.030)<br>0.025* |
| Multi-Unit<br>Housing | — | — | — | — | -0.012<br>(-0.021, -0.003)<br>0.009* |

## S6 Spatial random effect models

**Figure S9:**
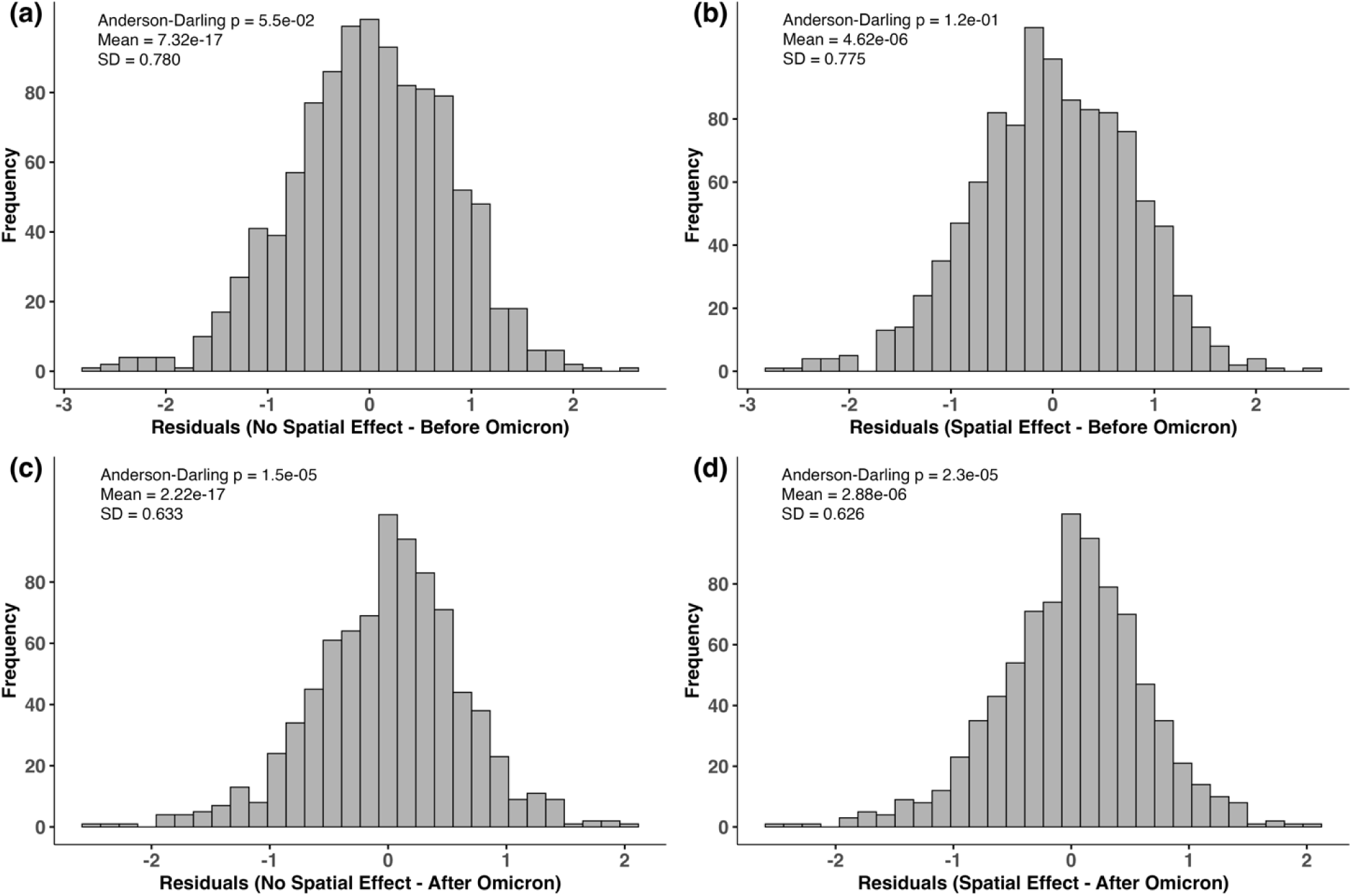
Comparison of residual distributions before and after incorporating spatial random effects in models fitted before and after Omicron dominance: (a) No spatial effect – before Omicron; (b) Spatial effect – before Omicron; (c) No spatial effect – after Omicron; (d) Spatial effect – after Omicron.

**Figure S10:**
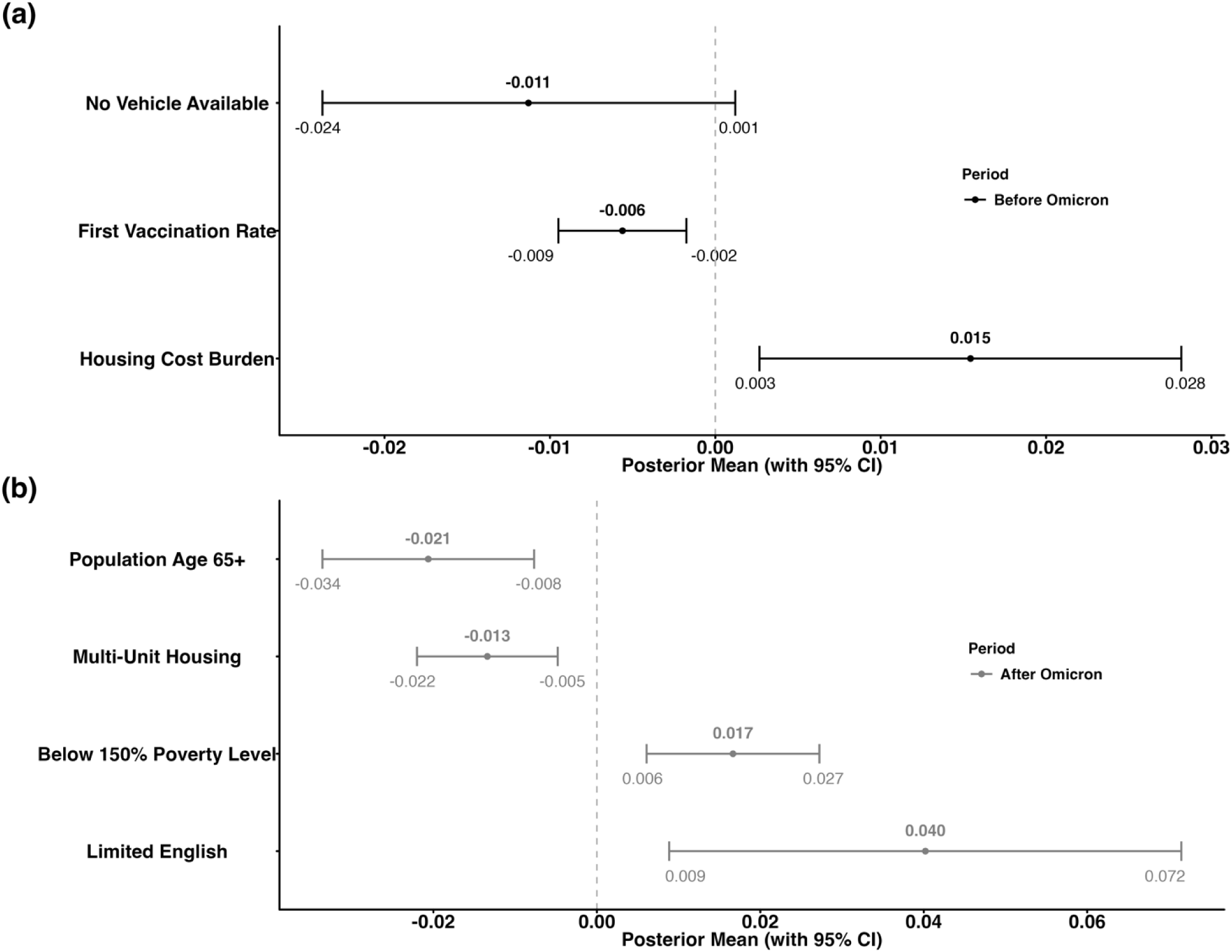
Spatial random effect models yielded posterior estimates (95% credible intervals (CIs)) of variables uniquely selected before (a) and after (b) Omicron dominance.

**Table S6.** Regional differences in long COVID incidence by social vulnerability factors thresholds.

| Variable | Region | Average incidence across counties (%) |  |  |  |
| --- | --- | --- | --- | --- | --- |
| | | Level ≤ Threshold | Level $>$ Threshold | Difference | p value |
| Disability Status Group<br>(threshold: 10%) | Midwest | 0.87 | 1.62 | 0.75 | <0.0001 |
|  | Northeast | 0.75 | 1.45 | 0.7 | <0.0001 |
|  | South | 0.61 | 1.46 | 0.85 | <0.0001 |
|  | Southwest | 1.33 | 1.13 | -0.2 | 0.0687 |
|  | West | 1.32 | 1.99 | 0.67 | 0.0007 |
| First Vaccine Rate<br>(threshold: 60%) | Midwest | 1.85 | 1.27 | -0.58 | <0.0001 |
|  | Northeast | 1.38 | 1.33 | -0.05 | 0.0254 |
|  | South | 1.75 | 1.2 | -0.55 | <0.0001 |
|  | Southwest | 1.26 | 0.99 | -0.27 | 0.0994 |
|  | West | 2.99 | 1.58 | -1.41 | <0.0001 |
| Group Quarters Population<br>(threshold: 3%) | Midwest | 1.48 | 1.71 | 0.23 | 0.0166 |
|  | Northeast | 1.27 | 1.41 | 0.14 | 0.0299 |
|  | South | 1.34 | 1.58 | 0.24 | <0.0001 |
|  | Southwest | 1.11 | 1.25 | 0.14 | 0.1059 |
|  | West | 1.89 | 1.82 | -0.07 | 0.9177 |
| Minority Population<br>(threshold: 20%) | Midwest | 1.61 | 1.37 | -0.24 | 0.0006 |
|  | Northeast | 1.6 | 0.91 | -0.69 | <0.0001 |
|  | South | 1.64 | 1.21 | -0.43 | <0.0001 |
|  | Southwest | 1.28 | 1.13 | -0.15 | 0.1022 |
|  | West | 2.22 | 1.59 | -0.63 | <0.0001 |
| Population Below 150%<br>Poverty<br>(threshold: 12%) | Midwest | 0.98 | 1.61 | 0.63 | <0.0001 |
|  | Northeast | 0.89 | 1.43 | 0.54 | 0.0003 |
|  | South | 0.7 | 1.46 | 0.76 | <0.0001 |
|  | Southwest | 1.69 | 1.13 | -0.56 | 0.0219 |
|  | West | 1.59 | 1.9 | 0.31 | 0.1979 |
| Rural-Urban Category<br>(Urban: Rural) | Midwest | 1.47 | 1.73 | 0.26 | <0.0001 |
|  | Northeast | 1.2 | 1.68 | 0.48 | <0.0001 |
|  | South | 1.17 | 1.98 | 0.81 | <0.0001 |
|  | Southwest | 1.03 | 1.65 | 0.62 | <0.0001 |
|  | West | 1.6 | 2.76 | 1.16 | <0.0001 |

